# Psychiatric symptoms, risk, and protective factors among university students in quarantine during the COVID-19 pandemic in China

**DOI:** 10.1101/2020.07.03.20144931

**Authors:** Shufang Sun, Simon B. Goldberg, Danhua Lin, Shan Qiao, Don Operario

**Affiliations:** Brown University Alpert Medical School, Department of Psychiatry and Human Behavior; University of Wisconsin-Madison College of Education, Department of Counseling Psychology; Beijing Normal University, Department of Psychology; University of South Carolina Arnold School of Public Health; Brown University School of Public Health, Department of Behavioral and Social Sciences

**Author notes:** Corresponding Author: Danhua Lin, Department of Psychology, Beijing Normal University, Beijing, China.

**Keywords:** COVID-19, psychiatric symptoms, stigma, trauma, young adult, China

## Abstract

This study investigated psychiatric symptoms (depression, anxiety, and traumatic stress) during state-enforced quarantine among university students in China. We conducted a cross-sectional survey with 1,912 university students during March and April 2020. Psychiatric symptoms in the mild or higher range based on clinical cut-offs were alarmingly prevalent: 67.05% reported traumatic stress symptoms, 46.55% had depressive symptoms, and 34.73% reported anxiety symptoms. Further, 19.56% endorsed some degree of suicidal ideation. We explored factors that may contribute to poor psychological health as well as those that may function as protective factors. Risk and protective factors examined included demographic variables, two known protective factors for mental health (mindfulness, perceived social support), four COVID-specific factors (COVID-19 related efficacy, perceived COVID-19 threat, perceived COVID-19 societal stigma, COVID-19 prosocial behavior) and screen media usage. Across psychiatric symptom domains, mindfulness was associated with lower symptom severity, while COVID-19 related financial stress, perceived COVID-19 societal stigma, and perceived COVID-19 threat were associated with higher symptom severity. COVID-19 threat and COVID-19 stigma showed main and interactive effects in predicting all mental health outcomes, with their combination associated with highest symptom severity. Average screen media device usage was 6 hours and usage was positively associated with depression. Female gender and COVID-19 prosocial behavior were associated with higher anxiety, while COVID-19 self-efficacy associated with lower anxiety symptoms. Study limitations and implications for treatment and prevention of affective disorders during crisis are discussed.

## Introduction

The 2019 novel coronavirus virus (COVID-19) pandemic, initially emerged in Wuhan, China in December 2019, has quickly become a major international public health crisis. In addition to the physical health consequences, public health authorities have expressed growing concerns regarding the psychological consequences of quarantine, social isolation, financial strain, and the threat of infection (Pfefferbaum & North, 2020; World Health Organization, 2020). The mental health impact of a pandemic of this scale is yet to be understood, and such an understanding is valuable for characterizing and ultimately addressing the psychological fallout of the current and future pandemics as well as increasing a basic scientific understanding of the psychiatric consequences of extreme stress (Holmes et al., 2020; Rauch, Simon, & Rothbaum, 2020; Sun, Lin, & Operario, 2020).

For several reasons, university students may be a particular vulnerable population to the mental health consequences of the COVID-19 pandemic. Developmentally, emerging adulthood is a vulnerable period: many mental disorders have symptoms that first occur during youth and young adulthood, which can negatively impact developmental trajectories through reduced educational achievement, increased substance use, and poor health behaviors (Arnett, Žukauskiene, & Sugimura, 2014; Patel, Flisher, Hetrick, & McGorry, 2007). The COVID-19 pandemic has led to massive disruption in the lives and education of university students in China, through school closure, and transition to internet-based learning, and social isolation from peers during state-enforced quarantine (Ministry of Education of China and Ministry of Industry and Information Technology of China, 2020). Available evidence suggests that these factors may indeed be leading to elevated psychiatric symptoms among university students in China. Two studies conducted in two different universities found rates of clinically elevated anxiety symptoms to be 15.4% (C. Wang & Zhao, 2020) and 24.9% (Cao et al., 2020) during the early outbreak of COVID-19. Another study with Chinese students from six universities in southwest China during February 2020 found 2.7% and 9.0% students reported moderate to severe forms of traumatic stress and depression, respectively (Tang et al., 2020). While highlighting high levels of distress, more research is needed to fully understand the psychological impact of the COVID-19 pandemic among university students in China. Beyond estimating prevalence, research identifying risk and protective factors is essential for increasing scientific understanding of the varied psychological reactions to large-scale infectious disease outbreaks and to guide intervention and prevention strategies.

The current study investigates the prevalence of mental health issues among university students in China during state-ordered quarantine and explores risk and protective factors. Three domains of psychiatric symptoms were surveyed, including depression, anxiety, and traumatic stress. Several demographic and contextual factors were explored as symptom correlates, including socioeconomic status, as COVID-19 has disproportionately impacted families from low-income backgrounds and financial hardship may affect mental health (Ahmed, Ahmed, Pissarides, & Stiglitz, 2020; Frasquilho et al., 2016). Two widely studied protective factors were explored, including mindfulness and perceived social support. Mindfulness, defined as “paying attention in a particular way, on purpose, in the present moment, and nonjudgmentally” (Kabat-Zinn, 1994, p. 4), has been consistently identified as a resilience factor for various types of psychological distress (Brown & Ryan, 2003; Goldberg et al., 2018; Keng, Smoski, & Robins, 2011). During the COVID-19 pandemic, mindfulness meditation has been actively promoted by Chinese health officials to the general public as a way to enhance psychological well-being (Zhang, Wu, Zhao, & Zhang, 2020). However, the role of mindfulness in the context of pandemic mental health has not been previously investigated. Perceived social support is another established protective factor for well-being (Bovier, Chamot, & Perneger, 2004; Hefner & Eisenberg, 2009), and it may be particularly crucial during the COVID-19 pandemic due to increased social isolation. Screen media usage was explored as a potentially relevant factor for the mental health of young adults in the pandemic context. Scholars have raised concerns regarding increased internet and smartphone addiction during the COVID-19 pandemic and the negative mental health impact of increased usage (Garfin, Silver, & Holman, 2020; Sun et al., 2020). However, to our knowledge, no empirical research has explored its role.

In addition, four COVID-19 related factors were explored as potential predictors, including COVID-19 prosocial behavior, COVID-19 self-efficacy, perceived COVID-19 threat, and perceived COVID-19 societal stigma. In general, altruistic and prosocial social behavior appears to promote well-being for the helpers (Curry et al., 2018; Martela & Ryan, 2016). From an evolutionary perspective, prosocial behavior in response to public health threats can be advantageous for both the group and individual (Kurzban, Burton-Chellew, & West, 2015). COVID-19 self-efficacy was explored as a predictor of mental health, as belief in one’s capacity to prevent COVID-19 and take necessary steps for treatment may facilitate an increased sense of control in an evolving outbreak (Person et al., 2004). Consistent with prior work showing perceived SARS threat predicted emotional exhaustion among frontline nurses during the SARS outbreak (Fiksenbaum, Marjanovic, Greenglass, & Coffey, 2006), we examined perceived COVID-19 threat (i.e., perception of one’s likelihood of contracting COVID-19) as a risk factor. Stigma was also examined as a potential factor related to mental health problems, as the early phase of an emerging outbreak tends to be characterized by intense disease-related stigma and fear due to its evolving nature and scientific uncertainties (Des Jarlais, Galea, Tracy, Tross, & Vlahov, 2006; Person et al., 2004). A recent study found 90% of respondents in China exhibited discriminatory attitudes toward people and regions associated with the COVID-19 outbreak (He, He, Zhou, Nie, & He, 2020). However, the role of societal stigma of an emerging infectious disease (i.e., perceived negative public attitude toward people and regions associated with the outbreak) on mental health outcomes of a general population is still unknown. Lastly, we examined potential interaction between perceived COVID-19 threat and perceived COVID-19 societal stigma. This was informed by the ecological model (person X context) (Bronfenbrenner, 1977; Cook, Purdie-Vaughns, Meyer, & Busch, 2014). Given the large-scale impact of COVID-19 reaching all individuals in the society, perceived COVID-19 societal stigma (a contextual, environmental-level factor) may amplify the association between perceived personal threat of COVID-19 and mental health outcomes.

We hypothesized that the following would function as risk factors, indicated by their positive association with psychiatric symptoms: COVID-related financial stress, screen media usage, perceived COVID-19 threat, perceived COVID-19 societal stigma. We also hypothesized an interactive relationship between perceived threat and perceived stigma, such that the link between perceived threat and psychiatric symptoms is increased in the presence of perceived stigma. We hypothesized the following as protective factors, indicated by their negative association with psychiatric symptoms: mindfulness, perceived social support, and COVID-19 prosocial behaviors.

## Methods

### Study Procedure

The study was approved by the Institutional Review Board at Beijing Normal University. Data were collected via an anonymous online survey. Recruitment took place online, through advertisement on popular websites and WeChat-based platforms targeting Chinese university students. Data collection occurred between March 20^th^ and April 10^th^ 2020, approximately two months following the official announcement of the COVID-19 outbreak in China (January 20^th^, 2020) while people were under state-enforced strict quarantine.

Inclusion criteria for participation included (a) being at least 18 years of age, (b) currently enrolled in a Chinese college or university as an undergraduate or graduate student, and (c) fluency in the Chinese language. Eligibility criteria and a consent form were provided on the survey’s welcome page. Participants were encouraged to take the survey on a personal device (e.g., computer, phone) in a private space. Participants received no monetary compensation. Following completion of the survey, participants were provided with suggestions for coping with psychological distress during the COVID-19 pandemic.

### Instruments

The survey included items assessing demographic information (e.g., age, gender, region, socioeconomic status). Participants were asked how much financial stress the COVID-19 pandemic has brought to their family, from 1 (*no financial stress*) to 5 (*significant financial stress*). A single-item question assessed participants’ screen media usage: in the past two weeks, outside of school and work time, *how many hours daily* on average have you spent on screen media (e.g., phone, computer)?

**Depression** was measured by the Patient Health Questionnaire-9 (PHQ-9; Chinese version), a depression screening tool (Kroenke, Spitzer, & Williams, 2001; W. Wang et al., 2014). PHQ-9 assesses symptoms of depression in the past two weeks. Each item reflects one of the nine DSM-IV criteria for major depressive episode (e.g., “little interest or pleasure in doing things,” “trouble concentrating on things, such as reading the newspaper or watching television”) (American Psychiatric Association, 1994). Participants indicate symptom severity from 0 (*not at all*) to 3 (*nearly every day*). Recommended clinical cut-off values are 5-9 (mild), 10-14 (moderate), 15-19 (moderately severe), and ≥20 (severe) (Kroenke et al., 2001). The scale has demonstrated good reliability and validity among the general population in China (W. Wang et al., 2014). Cronbach’s *α* was 0.93.

**Anxiety** symptoms were measured by the 7-item Generalized Anxiety Disorder Scale (GAD-7; Chinese version) (Spitzer, Kroenke, Williams, & Lowe, 2006; Zeng et al., 2013), a widely used screening tool for common anxiety disorders (e.g., Generalized Anxiety Disorder, Panic Disorder, Social Phobia). GAD-7 assesses symptoms of anxiety in the past two weeks (e.g., “feeling nervous, anxious, or on edge,” “feeling afraid as if something awful might happen”). Participants indicate symptom severity from 0 (*not at all*) to 3 (*nearly every day*). Recommended clinical cut-off values are 5-9 (mild), 10-14 (moderate), and ≥15 (severe) (Spitzer et al., 2006). The scale has demonstrated good reliability and validity in outpatient settings for people in China (Zeng et al., 2013). Cronbach’s *α* was 0.96.

**COVID-19 related traumatic stress** was assessed by the Impact of Events scale (IES; Chinese version) (Horowitz, Wilner, & Alvarez, 1979; Zhao, Wang, & Chang, 2003), a 15-item, widely used measure of event-specific distress. Participants were asked to indicate symptom severity in the context of the COVID-19 pandemic, from 0 (*not at all*) to 5 (*often*). Items assessed intrusion and avoidance clusters of posttraumatic stress disorder (PTSD) (e.g., “I tried not to talk about it,” “pictures about it popped into my mind”). Recommended clinical cut-off values are 9-25 (mild), 26-43 (moderate), and ≥44 (severe). The Chinese version has been used following natural disasters (e.g., earthquake) and has demonstrated good reliability and validity (L. Wang et al., 2011; Zhao et al., 2003). Cronbach’s *α* was 0.95.

**Mindfulness** was measured by the Chinese version of the Mindful Attention Awareness Scale (MAAS) (Brown & Ryan, 2003; Deng et al., 2012). The scale consists of 15 items and assesses dispositional mindfulness, namely receptive awareness of and attention to what is taking place in the present moment (e.g., “I find it difficult to stay focused on what’s happening in the present,”). Participants indicated frequency of each experience from 1 (*almost always*) to 6 (*almost never*), with a higher score indicating higher dispositional mindfulness. The MAAS has demonstrated good reliability and validity among people in China (Deng et al., 2012). Cronbach’s *α* was 0.94.

**Perceived social support** was measured by an adapted version of the Multidimensional Scale of Perceived Social Support (MSPSS)- Chinese version (Zimet, Dahlem, Zimet, & Farley, 1988). The original MSPSS has 12 items that assess perceived social support from three resources (family, friends, significant other). Considering that not all young adults are in a romantic relationship, only the eight items assessing perceived social support from family and friends were used (e.g., “my family really tries to help me,” “I can count on my friends when things go wrong”), which participants rated on a 5-point Likert scale (*1 = strongly disagree; 5 = strongly agree*). Cronbach’s *α* was 0.93.

**COVID-19 prosocial behavior** was assessed by items adapted from the Empathic Responding to SARS scale (Lee-Baggley, DeLongis, Voorhoeave, & Greenglass, 2004) and Prosocialness Scale (Caprara, Steca, Zelli, & Capanna, 2005). Participants rated nine items assessing prosocial behavior specific to COVID-19 (e.g., “I shared or posted scientific knowledge or uplifting news about COVID-19,” “I shared or donated resources (e.g., money, masks, medical gowns) to those in need”), ranging from 1 (*strongly agree*) to 5 (*strongly disagree*). Cronbach’s *α* was 0.93.

**COVID-19 Self-Efficacy** was measured through an adapted version of the Ebola-related Self-Efficacy scale (Cahyanto, Wiblishauser, Pennington-Gray, & Schroeder, 2016). Five items assessed perceived ability in adhering to COVID-19 prevention measures (e.g., “I am confident that I can understand health instructions about COVID-19 prevention”) rated as 1 (*strongly disagree*) to 4 (*strongly agree*). Cronbach’s *α* was 0.90.

**Perceived COVID-19 societal stigma** was measured via an adapted version of the Perceived External Stigma Subscale of the Ebola-related Stigma Questionnaire (Overholt et al., 2018), which itself was derived from Berger’s HIV Stigma Scale (Berger, Ferrans, & Lashley, 2001). Six items assessed perceived external, societal stigma against COVID-19, rated as 1 (*strongly disagree*) to 5 (*strongly agree*). Sample items include “most people are afraid of a person who has had COVID-19 or from regions severely affected by COVID-19”, “most people think that a person who has had COVID-19 is disgusting.” Higher total scores indicate higher levels of stigma. Cronbach’s *α* was 0.91.

**Perceived COVID-19 threat** was adapted from a measure previously used to assess Chinese people’s perceived threat by SARS (Lee-Baggley et al., 2004). Seven items assessed perceived infection threat by COVID-19 (e.g., “I don’t think I could get COVID-19”, “I think COVID-19 will threaten my health”) rated as 1 (*strongly disagree*) to 4 (*strongly agree*). Higher total scores indicate higher perceived COVID-19 threat. Cronbach’s *α* was 0.72.

### Data Analysis

First, bivariate analysis (correlation for continuous predictors, independent sample t-test for binary predictor) was conducted to identify variables that had significant associations with psychiatric symptoms (i.e., depression, anxiety, and traumatic stress). Second, variables identified in the first step as having significant bivariate associations with psychiatric symptoms were entered to a regression model simultaneously in order to identify significant factors on a multivariate level (Model 1). A second model for each outcome only included variables significant (*p* < .05) in model 1 (Model 2). This approach allows us to identify significant predictors, build a model with these factors (Model 2), and subsequently compare it with the regression model with the interaction term and controlled for significant covariates (Aiken & West, 1991; Cohen, Cohen, West, & Aiken, 2003). Third, to investigate the interactive effects of perceived COVID-19 threat and societal COVID-19 stigma, we conducted multiple regression analyses examining their interaction covarying significant predictors from Model 2 (Model 3). All continuous predictors were mean-centered in all models to avoid potential multicollinearity and for ease of understanding (i.e., a one-unit difference means one standard deviation difference). Gender as a binary variable was dummy coded (male = 0, female = 1). For models that yielded significant interaction effects, simple slopes analysis was employed to probe interactions at ±1 *SD* from the mean of the moderator (Aiken & West, 1991). Model comparison using ANOVA was conducted to further identify the proportion of variance explained by the interaction term.

## Results

### Descriptive and Bivariate Statistics

The sample included 1,912 Chinese young adult university students. Participants’ average age was 20.28 (*SD* = 2.10, Median = 20, Range = [18, 49]). The majority of participants were female (*n* = 1,334; 69.77%). Most participants (91.84%) were pursuing their undergraduate education. Participants resided in 30 out of the 36 provinces in China. Most participants noted financial stress on their family due to COVID-19, with the largest proportion (40.90%) reporting “some stress” (Table 1).

**Table 1.** Sample characteristics

| **Sociodemographic variables** | **N (%)** |
| --- | --- |
| **Gender** |  |
| Female | 1,334 (69.77) |
| Male | 578 (30.23) |
| **Socioeconomic status compared to classmates in school** |  |
| Lower than average | 675 (35.30) |
| On average | 1,166 (60.98) |
| Higher than average | 71 (3.71) |
| **Financial stress on family due to COVID-19** |  |
| Very significant stress | 70 (3.66) |
| Significant stress | 300 (15.69) |
| Some stress | 782 (40.90) |
| Little stress | 435 (22.75) |
| No stress | 325 (17.00) |
| **Education (currently pursuing)** |  |
| Undergraduate | 1,756 (91.84) |
| Master's education | 139 (7.27) |
| Doctorate education | 17 (0.89) |
| **Depression** |  |
| No symptoms (0-4) | 1,022 (53.45) |
| Mild symptoms (5-9) | 592 (30.96) |
| Moderate symptoms (10-14) | 169 (8.84) |
| Moderately severe (15-19) | 102 (5.33) |
| Severe symptoms (20-27) | 27 (1.41) |
| **Anxiety** |  |
| No symptoms (0-4) | 1,248 (65.27) |
| Mild symptoms (5-9) | 480 (25.10) |
| Moderate symptoms (10-14) | 158 (8.26) |
| Severe symptoms ($\geq 15$) | 26 (1.36) |
| **Traumatic stress** |  |
| No symptoms (0-8) | 630 (32.95) |
| Mild symptoms (9-25) | 944 (49.37) |
| Moderate symptoms (26-43) | 119 (6.22) |
| Severe symptoms ($\geq 44$) | 219 (11.45) |
| **Suicide ideation** |  |
| Never | 1,538 (80.44) |
| Rarely | 261 (13.65) |
| Sometimes | 93 (4.86) |
| Often | 20 (1.05) |
| **Region** |  |
| North China (Beijing, Tianjin, Hebei, Shanxi, Inner Mongolia) | 275 (14.38) |
| Northeast China (Liaoning, Jilin, Heilongjiang) | 61 (3.19) |
| East China (Shanghai, Jiangsu, Zhejiang, Anhui, Fujian, Jiangxi, Shandong) | 541 (28.29) |
| South Central China (Henan, Hubei, Hunan, Guangdong, Guangxi, Hainan) | 856 (44.77) |
| Southwest China (Chongqing, Sichuan, Guizhou, Yunnan, Tibet) | 109 (5.70) |
| Northwest China (Shaanxi, Gansu, Qinghai, Ningxia, Xinjiang) | 67 (3.50) |
| Hong Kong, Macau, or living abroad | 3 (0.16) |

Mental health symptoms were notably high. The majority of participants (67.05%) reported COVID-19-related traumatic stress symptoms within the clinical range (i.e., mild or higher). Depression symptoms were clinically elevated for 46.55% and anxiety symptoms for 34.73%. Approximately one in five (19.56%) reported some degree of suicidal ideation in the past two weeks (from rarely to often). Most clinical elevations were in the mild range. Proportion with moderate or higher clinical elevations were 17.67% for traumatic stress, 15.58% for depression, and 9.62% for anxiety.

Socioeconomic status, family financial stress due to COVID-19, mindfulness, perceived social support, COVID-19 self-efficacy, perceived COVID-19 societal stigma, and perceived COVID-19 threat had significant associations with all three outcomes (Table 2). Given that socioeconomic status and family financial stress due to COVID-19 were correlated (*r* = - .43, *p* < .001) and measured related constructs, only family financial stress due to COVID-19 was used as a predictor in the regression models. Age was weakly positively associated with anxiety. COVID-19 prosocial behavior and screen media usage were significantly associated with depression and anxiety. Anxiety symptoms were higher for female participants (*t* [1017] = -2.10, *p* = .036, mean [*M*] for male = 3.27, *M* for female = 3.74). Depressive symptoms and traumatic stress did not differ based on participants’ gender.

**Table 2.** Correlations among continuous variables

|  | Depression | Anxiety | Traumatic stress | Age | SES | Financial stress due to COVID-19 | Mindfulness | Social support | COVID-19 prosocial behavior | COVID-19 self-efficacy | Perceived COVID-19 stigma | Perceived COVID-19 threat | Screen media usage |
| --- | --- | --- | --- | --- | --- | --- | --- | --- | --- | --- | --- | --- | --- |
| Anxiety | .69*** | - |  |  |  |  |  |  |  |  |  |  |  |
| Traumatic stress | .47*** | .43*** | - |  |  |  |  |  |  |  |  |  |  |
| Age | .04 | .06** | .03 | - |  |  |  |  |  |  |  |  |  |
| SES | -.15*** | -.13*** | -.09*** | -.07** | - |  |  |  |  |  |  |  |  |
| Financial stress due to COVID-19 | .24*** | .24*** | .20*** | .08*** | -.43*** | - |  |  |  |  |  |  |  |
| Mindfulness | -.39*** | -.31*** | -.29*** | .02 | .12*** | -.20*** | - |  |  |  |  |  |  |
| Social support | -.30*** | -.26*** | -.11*** | .01 | .16*** | -.17*** | .28*** | - |  |  |  |  |  |
| COVID-19 prosocial behavior | -.13*** | -.10*** | .01 | .03 | .10*** | -.06* | .21*** | .47*** | - |  |  |  |  |
| COVID-19 self-efficacy | -.18*** | -.20*** | -.10*** | .01 | .12*** | -.14*** | .20*** | .43*** | .36*** | - |  |  |  |
| Perceived COVID-19 stigma | .37*** | .32*** | .42*** | -.01 | -.08*** | .17*** | -.24*** | -.22*** | -.14*** | -.23*** | - |  |  |
| Perceived COVID-19 threat | .18*** | .22*** | .20*** | -.08*** | -.04 | .09*** | -.008*** | -.01 | .06*** | .02 | .14*** | - |  |
| Screen media usage | .11*** | .06* | .04 | -.10*** | -.04 | .03 | -.10*** | -.09*** | -.07** | -.02 | .02 | .01 | - |
| *M* | 5.20 | 3.59 | 19.83 | 20.38 | 1.68 | 2.66 | 67.14 | 32.40 | 26.70 | 16.44 | 11.94 | 17.68 | 6.18 |
| *SD* | 5.33 | 4.35 | 15.98 | 2.10 | 0.54 | 1.05 | 13.27 | 5.86 | 5.61 | 2.50 | 3.98 | 3.52 | 3.05 |

Note. $p < .05$ \*, $p < .01$ \*\*, $p < .001$ \*\*\*

### Predicting Depressive Symptoms

COVID-19 prosocial behavior and COVID-19 self-efficacy were no longer significant predictors when all identified significant variables were entered to the regression model simultaneously (Model 1). The final model without interaction (Model 2; Table 3) included six significant predictors of depressive symptoms. Risk factors (i.e., positive correlates) for depressive symptoms included financial stress due to COVID-19 (*β* = 0.119), two COVID-specific risk factors (perceived COVID-19 threat: *β* = 0.109 perceived COVID-19 societal stigma: *β* = 0.238), and screen media device usage (*β* = 0.058). Protective factors (i.e., negative correlates) for depressive symptoms included mindfulness (*β* = -0.249) and perceived social support (*β* = -0.153). The full model explained 28.2% of variance in depression (i.e., adjusted *R*^2^ = .282).

**Table 3.**
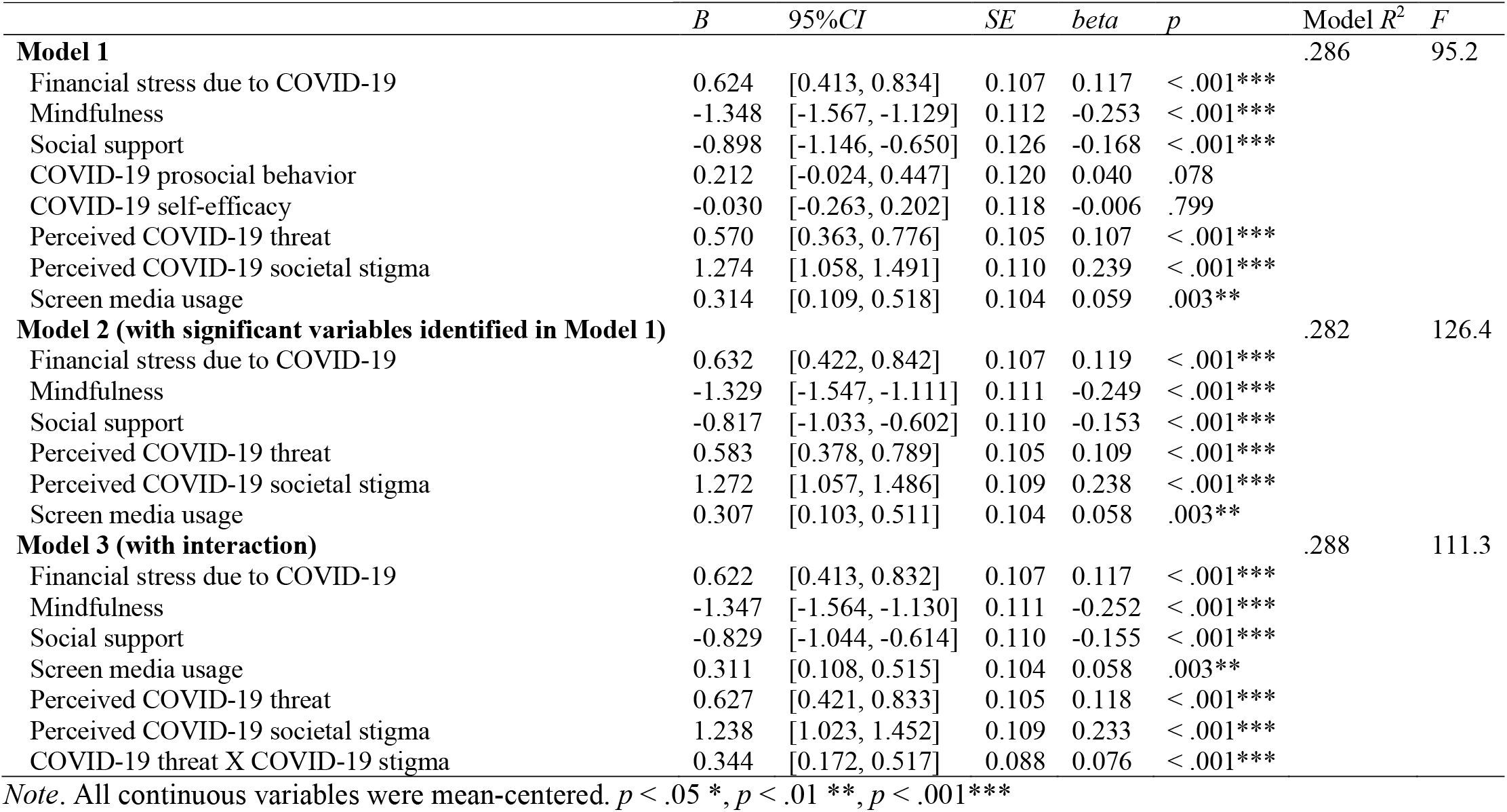
Predictors of depressive symptoms

A significant interaction was detected between perceived COVID-19 threat and perceived COVID-19 societal stigma (Model 3, *β* = 0.076, *p* < .001), after accounting for all other significant variables identified in Model 2. As predicted, the positive association between perceived COVID-19 threat with depressive symptoms was amplified in the presence of higher levels of perceived COVID-19 societal stigma. Model comparison through analysis of covariance (ANCOVA) indicated superiority of Model 3 to Model 2, *F* = 15.38, *p* < .001, and the interaction term accounted for 6% additional variance. Simple slope analyses indicated that perceived COVID-19 threat remained a significant predictor of depression when perceived COVID-19 societal stigma was 1 *SD* above (*β* = 0.182 [0.129, 0.235], *p* < .001) or below the mean (*β* = 0.053 [0.005, 0.101], *p* = .029), although depression was notably higher when perceived COVID-19 societal stigma was increased (Figure 1a).

**Figure 1.**
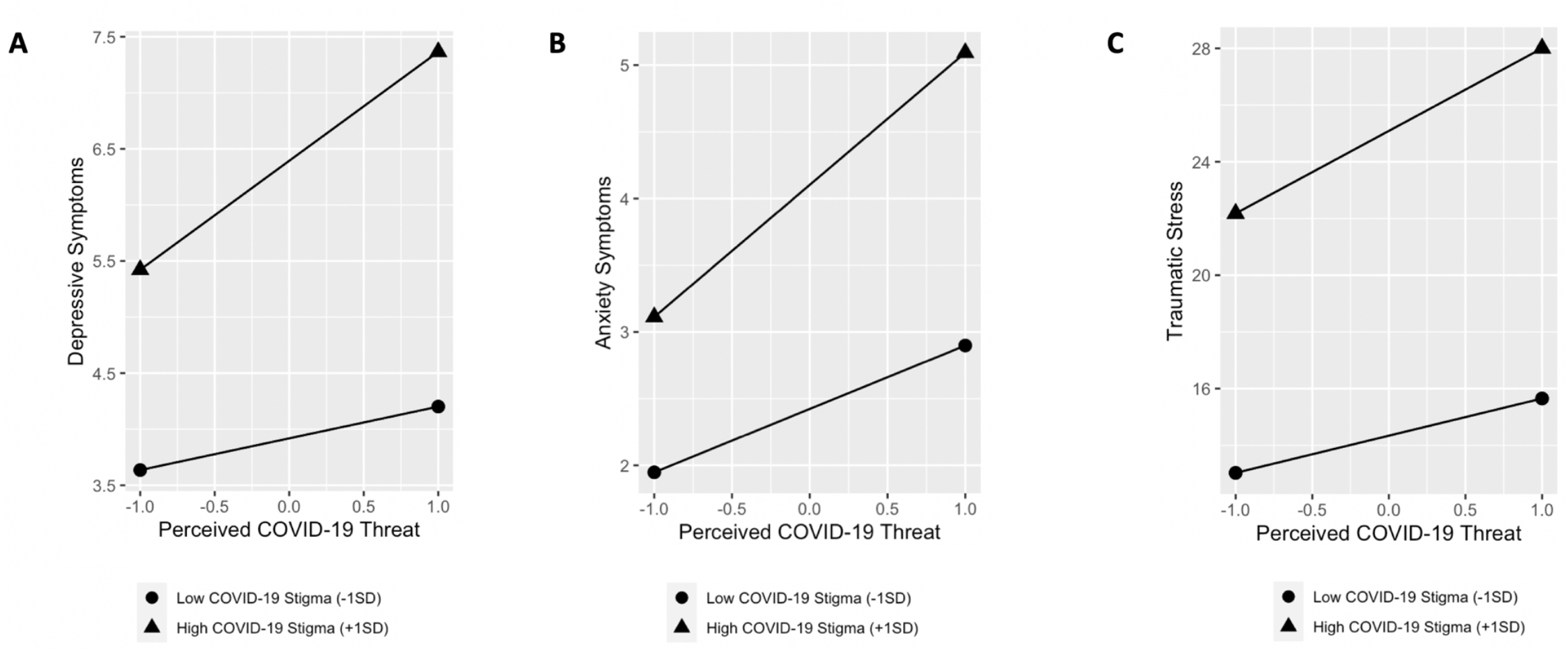
Perceived COVID-19 threat and perceived COVID-19 societal stigma interact when predicting psychiatric symptoms

### Predicting Anxiety Symptoms

Screen media device usage was no longer a significant predictor in the regression model (Table 4; Model 1). The final model without interaction term (Model 2) included three demographic variables (age: *β* = 0.073, gender: *β* = 0.045, financial stress due to COVID-19: *β* = 0.124), two known protective factors for anxiety symptoms including mindfulness (*β* = -0.190) and social support (*β* = -0.143), and four COVID-19 specific factors including COVID-19 prosocial behavior (*β* = 0.049), COVID-19 self-efficacy (*β* = -0.055), perceived COVID-19 threat (*β* = 0.160), and perceived COVID-19 societal stigma (*β* = 0.199). Contrary to hypothesis, higher level of COVID-19 prosocial behavior positively associated with more anxiety symptoms.

**Table 4.** Predictors of anxiety symptoms

|  | *B* | *95%CI* | *SE* | *beta* | *p* | Model *R*<sup>2</sup> | *F* |
| --- | --- | --- | --- | --- | --- | --- | --- |
| **Model 1** |  |  |  |  |  | .236 | 59.9 |
| Age | 0.327 | [0.154, 0.501] | 0.089 | 0.075 | < .001*** |  |  |
| Gender | 0.416 | [0.039, 0.792] | 0.192 | 0.044 | .030* |  |  |
| Financial stress due to COVID-19 | 0.538 | [0.360, 0.716] | 0.091 | 0.124 | < .001*** |  |  |
| Mindfulness | -0.818 | [-1.003, -0.634] | 0.094 | -0.188 | < .001*** |  |  |
| Social support | -0.614 | [-0.827, -0.404] | 0.107 | -0.141 | < .001*** |  |  |
| COVID-19 prosocial behavior | 0.216 | [0.018, 0.415] | 0.101 | 0.050 | .033* |  |  |
| COVID-19 self-efficacy | -0.244 | [-0.440, -0.048] | 0.100 | -0.056 | < .001*** |  |  |
| Perceived COVID-19 threat | 0.699 | [0.524, 0.873] | 0.089 | 0.161 | < .001*** |  |  |
| Perceived COVID-19 societal stigma | 0.867 | [0.684, 1.049] | 0.093 | 0.199 | < .001*** |  |  |
| Screen media usage | 0.109 | [-0.064, 0.282] | 0.088 | 0.025 | .218 |  |  |
| **Model 2 (with significant variables identified in Model 1)** |  |  |  |  |  | .235 | 66.4 |
| Age | 0.317 | [0.144, 0.490] | 0.088 | 0.073 | < .001*** |  |  |
| Gender | 0.423 | [0.047, 0.800] | 0.192 | 0.045 | .027* |  |  |
| Financial stress due to COVID-19 | 0.539 | [0.362, 0.717] | 0.091 | 0.124 | < .001*** |  |  |
| Mindfulness | -0.827 | [-1.011, -0.643] | 0.094 | -0.190 | < .001*** |  |  |
| Social support | -0.621 | [-0.830, -0.412] | 0.106 | -0.143 | < .001*** |  |  |
| COVID-19 prosocial behavior | 0.213 | [0.014, 0.412] | 0.101 | 0.049 | .0236* |  |  |
| COVID-19 self-efficacy | -0.240 | [-0.436, -0.044] | 0.100 | -0.055 | .016* |  |  |
| Perceived COVID-19 threat | 0.698 | [0.523, 0.873] | 0.089 | 0.160 | < .001*** |  |  |
| Perceived COVID-19 societal stigma | 0.866 | [0.683, 1.048] | 0.093 | 0.199 | < .001*** |  |  |
| **Model 3 (with interaction)** |  |  |  |  |  | .240 | 61.3 |
| Age | 0.325 | [0.152, 0.497] | 0.088 | 0.075 | < .001*** |  |  |
| Gender | 0.424 | [0.049, 0.799] | 0.191 | 0.045 | .027* |  |  |
| Financial stress due to COVID-19 | 0.532 | [0.355, 0.710] | 0.090 | 0.122 | < .001*** |  |  |
| Mindfulness | -0.839 | [-1.022, -0.655] | 0.094 | -0.193 | < .001*** |  |  |
| Social support | -0.623 | [-0.832, -0.415] | 0.106 | -0.143 | < .001*** |  |  |
| COVID-19 prosocial behavior | 0.196 | [-0.002, 0.394] | 0.101 | 0.045 | .053 |  |  |
| COVID-19 self-efficacy | -0.238 | [-0.433, -0.043] | 0.099 | -0.055 | .017* |  |  |
| Perceived COVID-19 threat | 0.733 | [0.558, 0.909] | 0.089 | 0.169 | < .001*** |  |  |
| Perceived COVID-19 societal stigma | 0.840 | [0.658, 1.022] | 0.093 | 0.193 | < .001*** |  |  |
| COVID-19 threat X COVID-19 stigma | 0.258 | [0.112, 0.403] | 0.074 | 0.070 | < .001** |  |  |

Note. All continuous variables were mean-centered. $p < .05$ \*, $p < .01$ \*\*, $p < .001$ \*\*\*

Accounting for all significant variables identified in Model 2, the interaction term of perceived COVID-19 threat X perceived COVID-19 societal stigma was significant in predicting anxiety symptoms (Table 4; Model 3), *β* = 0.070, *p* < .001. Main effects of perceived COVID-19 threat and perceived COVID-19 societal stigma remained significant. Similarly, higher levels of stigma further intensified the relationship between perceived COVID-19 threat and anxiety symptoms. Model 3 was superior to model 2, *F* = 12.09, *p* < .001, accounting for 5% more variance. Simple slope analysis found that high levels of perceived COVID-19 societal stigma intensified the positive association between perceived COVID-19 threat and anxiety symptoms (*β* = 0.228 [0.173, 0.283], *p* < .001; Figure 1b). At 1 *SD* below the mean of perceived COVID-19 societal stigma, it still heightened the harmful effect of perceived COVID-19 threat on anxiety symptoms, *β* = 0.109 [0.060, 0.159], *p* < .001.

### Predicting Traumatic Stress

COVID-19 related traumatic stress was significantly predicted by financial stress due to COVID-19 (*β* = 0.100), mindfulness (*β* = -0.178), perceived COVID-19 threat (*β* = 0.136), and perceived COVID-19 societal stigma (*β* = 0.341) (Table 5; Model 2). Model’s adjusted *R*^2^ = 0.238, *F* = 150.0, *p* < .001 (Table 5; Model 2).

**Table 5.** Predictors of traumatic stress

|  | *B* | *95%CI* | *SE* | *beta* | *p* | Model *R*<sup>2</sup> | *F* |
| --- | --- | --- | --- | --- | --- | --- | --- |
| **Model 1** |  |  |  |  |  | .238 | 100.7 |
| Financial stress due to COVID-19 | 1.675 | [1.026, 2.323] | 0.331 | 0.105 | < .001*** |  |  |
| Mindfulness | -2.995 | [-3.666, -2.324] | 0.342 | -0.187 | < .001*** |  |  |
| Social support | 0.602 | [-0.114, 1.318] | 0.365 | 0.038 | .100 |  |  |
| COVID-19 self-efficacy | 0.123 | [-0.581, 0.827] | 0.356 | 0.008 | .732 |  |  |
| Perceived COVID-19 threat | 1.976 | [1.341, 2.612] | 0.324 | 0.124 | < .001*** |  |  |
| Perceived COVID-19 societal stigma | 5.570 | [4.902, 6.238] | 0.341 | 0.349 | < .001*** |  |  |
| **Model 2 (with significant variables identified in Model 1)** |  |  |  |  |  | .238 | 150.0 |
| Financial stress due to COVID-19 | 1.604 | [0.959, 2.249] | 0.329 | 0.100 | < .001*** |  |  |
| Mindfulness | -2.844 | [-3.498, -2.190] | 0.333 | -0.178 | < .001*** |  |  |
| Perceived COVID-19 threat | 2.011 | [1.377, 2.645] | 0.323 | 0.136 | < .001*** |  |  |
| Perceived COVID-19 societal stigma | 5.450 | [4.797, 6.104] | 0.333 | 0.341 | < .001*** |  |  |
| **Model 3 (with interaction)** |  |  |  |  |  | .241 | 122.2 |
| Financial stress due to COVID-19 | 1.584 | [0.941, 2.228] | 0.328 | 0.099 | < .001*** |  |  |
| Mindfulness | -2.892 | [-3.546, -2.239] | 0.333 | -0.181 | < .001*** |  |  |
| Perceived COVID-19 threat | 2.113 | [1.476, 2.749] | 0.324 | 0.132 | < .001*** |  |  |
| Perceived COVID-19 societal stigma | 5.375 | [4.721, 6.029] | 0.335 | 0.336 | < .001*** |  |  |
| COVID-19 threat X COVID-19 stigma | 0.801 | [0.268, 1.333] | 0.271 | 0.059 | .003* |  |  |

*Note.* All continuous variables were mean-centered. $p < .05$ \*, $p < .01$ \*\*, $p < .001$ \*\*\*

Accounting for significant variables identified in Model 2, as well as main effects of perceived COVID-19 threat and perceived COVID-19 societal stigma, their interaction term significantly predicted traumatic stress, *β* = 0.059. The model accounted for 24.1% of the variance in traumatic stress, *F* = 122.2, *p* < .001. Model comparison via ANOVA indicated that model 3 with interaction terms was superior to model 2, *F* = 8.70, *p* = .003, and accounted for 3% additional variance. Simple slope analysis revealed similar patterns: high levels of perceived COVID-19 societal stigma intensified the positive association between perceived COVID-19 threat and traumatic stress (*β* = 0.182 [0.128, 0.237], *p* < .001; Figure 1c). At 1 *SD* below the mean of perceived COVID-19 societal stigma, it still heightened the positive association between perceived COVID-19 threat and traumatic stress, though to a less degree (*β* = 0.082 [0.033, 0.131], *p* = .001).

Variance inflation factor (VIF) analysis across models predicting all domains of symptoms did not suggest multicollinearity (VIF values ranged from 1.02 to 1.50).

## Discussion

This large-scale, cross-sectional study investigated psychiatric symptoms among Chinese university students in strict quarantine during the early months of the COVID-19 outbreak. Our results are largely consistent with prior studies in China and elsewhere highlighting the adverse psychological consequences in the general population of both quarantine and disease outbreak (Brooks et al., 2020; Cao et al., 2020; Tang et al., 2020). This study extends prior work by highlighting several risk and protective factors related to mental health in university students. Across symptoms domains (depression, anxiety, traumatic stress), COVID-related financial stress, perceived threat, and perceived societal stigma emerged as consistent positive correlates of psychopathology. Perceived COVID-19 threat and COVID-19 societal stigma showed synergistic interaction, such that the presence of both were associated with additive effects when predicting all three symptom domains, highlighting the importance of contextual factors (i.e., ecological perspective; Bronfenbrenner, 1977). Both mindfulness and perceived social support emerged as protective factors, with mindfulness negatively associated with all three forms of psychopathology and perceived social support negatively associated with depression and anxiety. Screen media usage was weakly associated with depressive symptoms.

The prevalence on depression, anxiety, and traumatic stress among university students uncovered from this study is similar yet slightly higher than findings reported in two other recent studies concerning the initial phase of the outbreak (Cao et al., 2020; Tang et al., 2020). The prevalence of clinically elevated depressive symptoms in our sample (46.6%) is roughly twice as high as meta-analytic estimates of clinical elevations among Chinese college students prior to the pandemic (23.8%) (Lei, Xiao, Liu, & Li, 2016). To our knowledge, no meta-analyses or nationally representative survey data are available estimating rates of clinically elevated anxiety or traumatic stress symptoms among Chinese university students. Notably, while clinical elevations were fairly common in our sample, most participants scored in the mild range. Nonetheless, prevalence rates indicate continued high mental health needs within this population during the two months of quarantine period following the initial outbreak.

The COVID-19 pandemic has evolved into a serious threat to public health given its geographic reach, prolonged impact, and lack of cure or effective treatment. Thus, it is not surprising that perceived COVID-19 threat, namely the felt sense of threat to one’s health and life by COVID-19, predicted elevated psychiatric symptoms. Public health interventions may need to balance strategies to increase preventive behaviors against COVID-19 (e.g., social distancing, wearing masks) while attending to potentially detrimental mental health effects due to an increased sense of COVID-19 threat.

The significant role of perceived societal stigma on mental health, both alone and in interaction with perceived threat, is a novel and potentially important finding. Research on infectious disease stigma has focused on those infected or most vulnerable to infection (e.g., anticipated HIV stigma among men who have sex with men) (Stangl, Lloyd, Brady, Holland, & Baral, 2013). However, our study found that in the context of COVID-19, perceived societal stigma adversely affects the mental health of the general public (i.e., university students, not a group particularly vulnerable for infection or death). The mechanism through which perceived COVID-19 societal stigma operates to effect mental health is unclear; it might be related to the potential deteriorating impact of societal stigma on people’s collective self-esteem and societal belonging, as well as increased survivor guilt and empathic concerns for those affected in a high stigma environment (O’Connor, Berry, Weiss, & Gilbert, 2002; Taylor, 2019). Further, perceived COVID-19 stigma and perceived COVID-19 threat had a synergetic effect on all mental health outcomes, supporting an ecological perspective (person X context) (Bronfenbrenner, 1977; Cook et al., 2014) in understanding and addressing consequences of stigma. Therefore, interventions aimed at lowering public stigma could help reduce the psychological consequences of the COVID-19 pandemic.

As the economic consequences of COVID-19 have continued to unfold, it has become clear that individuals from lower socioeconomic background have been disproportionately burdened (Ahmed et al., 2020; Buheji et al., 2020). In keeping, we found that family financial stress due to COVID-19 consistently predicted psychiatric symptoms. This relationship is not new; past economic recessions have witnessed increased rates of common mental disorders and suicidal behavior in the general population (Frasquilho et al., 2016; Nordt, Warnke, Seifritz, & Kawohl, 2015; Reeves et al., 2012). In the context of the COVID-19 pandemic, economically impacted individuals may face the dual burden of COVID-19 threat and financial stress— both of which may adversely impact mental health. In addition to financial stress, being older and female were also identified as risk factors of anxiety symptoms. Students who are older may experience more pandemic-related stress related to employment and career development. Female students reported greater levels of anxiety, which is consistent with existing evidence that women report greater anxiety and more likely to develop anxiety disorders than men (McLean & Anderson, 2009).

Mindfulness was identified as a protective factor across all three symptoms domains. In the midst of great uncertainty, anxiety, and despair during the COVID-19 pandemic, dispositional attentiveness to the present moment may protect young adults from excessive worries, rumination, and fear. This is supported by evidence suggesting decreased rumination may be one of the key mechanisms underlying the efficacy of mindfulness-based interventions (Gu, Strauss, Bond, & Cavanagh, 2015). Given potentially increased demand and decreased availability of psychological services during the pandemic, evidence-based interventions that can enhance mindfulness may be a promising approach for mental health promotion (e.g., mindfulness delivered via mobile health; Linardon, 2020). Perceived social support also emerged as a protective factor for depression and anxiety. Loneliness and social isolation from peers may contribute to heightened distress for young adults during quarantine. Interventions that can safely enhance social connection (e.g., while maintaining social distance) are warranted.

Excessive screen time has been linked to a variety of health concerns including obesity, sleep disturbance, and mental health issues (Boone, Gordon-Larsen, Adair, & Popkin, 2007; Feng, Zhang, Du, Ye, & He, 2014). In our sample, participants’ self-reported an average of six hours of daily screen media time outside of school and work purposes in the past two weeks. In the context of the pandemic, young adults may consume more screen media due to restricted access to other avenues of entertainment and increased media exposure related to COVID-19 (e.g., news, report). Screen media usage weakly predicted depression but not other symptoms. It is possible that screen media engagement may have a mixed role during the pandemic. For instance, videoconferencing with family and friends could enhance social support while excessive TV watching by oneself may increase risk.

### Limitations

The current study has several limitations. First, although the current study recruited a geographically diverse national sample compared to previous studies on the mental health of Chinese university students restricted to a few universities (Cao et al., 2020; Tang et al., 2020; C. Wang & Zhao, 2020), the open recruitment method via the internet has its drawbacks. In particular, those highly impacted by COVID-19 may be particularly interested in enrollment and participation, which could upwardly bias estimates of psychiatric symptom severity. Thus, findings may not be representative of the larger Chinese university student population. Second, given the cross-sectional nature of the study, causal directions of the observed relationships cannot be ascertained. For instance, contrary to hypotheses, COVID-19 prosocial behavior was associated with heightened anxiety, and this could be due to those who experience higher anxiety during the pandemic being more likely to engage in prosocial actions as a coping approach. It is also likely that those prone to psychiatric symptoms (e.g., with pre-existing depression or anxiety disorders) may perceive COVID-19 as more threatening. It is crucial that future longitudinal and experimental studies further explore these associations. Third, the Impact of Events Scale (IES) (Horowitz et al., 1979) only measured two clusters of PTSD symptoms (intrusion and avoidance), missing negative alternations in cognition or mood and hyperarousal symptoms. Fourth, the PHQ-9 and GAD-7 are brief measures designed to indicate possible psychopathology. It would have been preferable to include longer self-report measures or clinician-rated measures to more accurately define prevalence rates. Similarly, given the self-report method, all variables are at risk to known biases (e.g., social desirability).

## Conclusion

Assessing psychiatric symptom prevalence and identifying risk and protective factors during the COVID-19 outbreak are critical to understanding and addressing the short-term and long-term psychological consequences of the COVID-19 pandemic. In addition to informing current and future pandemic responses, this research can also clarify the psychological consequences, risk, and protective factors of acute stress more generally. Conducted in March and April 2020 during massive, state-enforced quarantine, the current study found high rates of psychiatric symptoms in a sample of internet-recruited university students in China. Findings suggest high need for psychological health promotion among university students. Interventions targeting multi-level factors, including promoting mindfulness and social support at individual and interpersonal levels while reducing societal stigma, may be particularly promising. Attending to needs of disadvantaged groups most financially vulnerable may require both psychological and economic interventions.

## Data Availability

Data will be made available upon request.

